# Epigenomic profiling of the infrapatellar fat pad in osteoarthritis

**DOI:** 10.1101/2023.05.22.23290348

**Authors:** Peter Kreitmaier, Young-Chan Park, Diane Swift, Arthur Gilly, J Mark Wilkinson, Eleftheria Zeggini

**Affiliations:** Technical University of Munich (TUM) and Klinikum Rechts der Isar, TUM School of Medicine, Ismaninger Str. 22, 81675 Munich, Germany; Graduate School of Experimental Medicine, TUM School of Medicine, Technical University of Munich, 81675 Munich, Germany; Institute of Translational Genomics, Helmholtz Zentrum München, German Research Center for Environmental Health, 85764 Neuherberg, Germany; Department of Oncology and Metabolism, The University of Sheffield, Sheffield S10 2RX, UK

**Author notes:** joint corresponding author Correspondence.

**Keywords:** DNA methylation, osteoarthritis, infrapatellar fat pad, methylation QTL, EWAS

## Abstract

Osteoarthritis is a prevalent, complex disease of the joints, and affects multiple intra-articular tissues. Here, we have examined genome-wide DNA methylation profiles of primary infrapatellar fat pad and matched blood samples from 70 osteoarthritis patients undergoing total knee replacement surgery. Comparing the DNA methylation profiles between these tissues reveal widespread epigenetic differences. We produce the first genome-wide methylation quantitative trait locus (mQTL) map of fat pad, and make the resource available to the wider community. Using two-sample Mendelian randomization and colocalization analyses, we resolve osteoarthritis GWAS signals and provide insights into the molecular mechanisms underpinning disease aetiopathology. Our findings provide the first view of the epigenetic landscape of infrapatellar fat pad primary tissue in osteoarthritis.

## Introduction

Osteoarthritis is a complex joint disease that affects more than 300 million people^1^. In the face of aging populations, the impact of osteoarthritis on public health systems is estimated to increase further^1^. Current treatment methods are limited to pain management and total joint replacement, highlighting the need to develop novel, personalised treatment strategies. Therefore, it is important to enhance our understanding of the genetic and genomic basis of osteoarthritis.

To date, genome-wide association analyses (GWAS) have identified more than 150 genetic risk loci^2^ of osteoarthritis, thus improving our understanding of its polygenic basis. Large-scale molecular datasets of relevant, primary cell types of osteoarthritis patients can reveal molecular mechanisms underlying disease and provide insights beyond genetic studies. Combining results from genetic and molecular studies can help pinpoint molecular mechanisms of disease development and progression, specifically the likely effector genes through which genetic risk variants exert their effect on osteoarthritis development in affected tissues.

Whilst a number of studies have investigated genome-wide molecular profiles of osteoarthritis-affected primary joint tissues^3,4^ the majority have focused on cartilage^5^. Osteoarthritis affects all joint tissues, and a small number of genome-wide molecular studies have extended molecular profiling to other primary joint tissues, such as the synovium^6,7^ or subchondral bone^8^.

The infrapatellar fat pad, an adipocyte-rich tissue located inferior to the patella in the anterior part of the knee joint^9^, has not been deeply studied in osteoarthritis to date. The fat pad is located among other joint tissues and protects knee components (by stabilising the patella) when exposed to mechanical stress, e.g. during exercise. In osteoarthritis-affected knees, the infrapatellar fat pad undergoes disease-related alterations, including fibrosis, inflammation and vascularization. Furthermore, it is traversed by nerves and therefore constitutes a source of knee osteoarthritis-related pain.

The fat pad may also interact with other joint tissues during osteoarthritis development and progression ^9^. For example, it is proposed that the fat pad secretes pro-inflammatory and catabolic factors that promote cartilage degeneration and inhibit repair mechanisms^10^. Studies using chondrocyte cultures and fat pad-derived fat-conditioned media have provided some first insights into the potential cross-talk between the fat pad and cartilage^9^.

Furthermore, the fat pad lies adjacent to the synovium, a connective tissue that lines the joint capsule. Both tissues undergo similar osteoarthritis-related changes, e.g. develop a similar immune cell profile^11^. Studies *in vitro* and in mouse models suggest interactions between these tissues ^12–14^. For example, Bastiaansen-Jenniskens et al. cultured fibroblast-like synoviocytes in fat-conditioned medium from fat pad samples of knee osteoarthritis patients, and suggest that fat pad induces fibrotic changes in synoviocytes by stimulating collagen synthesis as well as cell proliferation and migration^14^.

Only a small number of studies have examined the profile of infrapatellar fat pad in osteoarthritis patients. Gandhi et al. characterised microarray-based gene expression profiles of the infrapatellar fat pad in 34 (29 and five in late and early stage knee osteoarthritis, respectively) individuals ^15^. Other studies have investigated the molecular characteristics of osteoarthritis fat pad in genomic regions of osteoarthritis risk signals ^16–18^ or focused on cytokines and extracellular matrix genes ^19^.

In this study, we focus on DNA methylation, an epigenetic mark that refers to the covalent addition of a methyl-group to the DNA. Methylation is dynamic, tissue-specific, and plays a regulatory role in gene expression. In general, promoter methylation is negatively correlated with gene expression, whereas methylation in other parts of the genome, such as the gene body, remain less understood.

We examine the genome-wide DNA methylation profile of infrapatellar fat pad adipocytes of osteoarthritis-affected knees. We (1) compare fat pad and blood methylation profiles matched from the same patients, (2) generate a genome-wide methylation quantitative trait loci (mQTL) map in fat pad and (3) resolve osteoarthritis GWAS signals by integrating omics with genetic association data.

## Subjects and methods

### Study participants

We have collected tissue samples from 210 patients undergoing total joint replacement surgery (111 women, 99 men, age 48-93 years, mean 71 years). All patients provided written, informed consent prior to participation in the study. Adipose tissue was collected from the infra-patellar fat pad by sharp dissection of the fat tissue from the surface of the patellar ligament to yield not less than 1cm^3^ of homogeneous adipose tissue. This work was approved by Oxford NHS REC C (10/H0606/20, SC/15/0132 and SC/20/0144), and samples were collected under Human Tissue Authority license 12182, South Yorkshire and North Derbyshire Musculoskeletal Biobank, University of Sheffield, UK. We confirmed a joint replacement for osteoarthritis, with no history of significant knee surgery (apart from meniscectomy), knee infection, or fracture, and no malignancy within the previous 5 years. We further confirmed that no patient used glucocorticoid use (systemic or intra-articular) within the previous 6 months, or any other drug associated with immune modulation. We also obtained a peripheral blood sample to extract DNA from all patients.

### Adipocyte and peripheral blood collection and processing

Adipose tissue samples were transported in Dulbecco’s modified Eagle’s medium (DMEM)/F-12 (1:1) (Life Technologies) supplemented with 2 mM glutamine (LifeTechnologies), 100 U/ml penicillin, 100 μg/ml streptomycin (Life Technologies),2.5 μg/ml amphotericin B (Sigma-Aldrich) and 50μg/ml ascorbic acid (Sigma-Aldrich) (serum free media). Next, the adipose tissue samples were cut into small pieces (<2mm^3^) and digested in 3 mg/ml collagenase type I (Sigma-Aldrich) in serum free media for 1 hour at 37 °C on a flatbed shaker and resuspended in 2mls of PBS and passed through a 100μm cell strainer (Fisher Scientific). Next, the eluent was made up to 10mls in PBS and centrifuged at 23g for 5 min. Subsequently, the cell pellet was washed twice in PBS and centrifuged at 323g for a further 5 mins. Cells were counted using a haemocytometer and the viability checked using trypan blue exclusion (Invitrogen). The resulting cell pellet was resuspended in 650μl of RLT buffer (Qiagen) and DTT Dithiothreitol (20ul DTT per 1ml of RLT). The optimal cell number for spin column extraction from cells was between 4 × 10^6^ and 1 × 10^7^. Cells were then pelleted and homogenised. DNA extraction was carried out using Qiagen AllPrep DNA Mini Kit following the manufacturer’s instructions. Samples were flash frozen in liquid nitrogen and stored at −80°C prior to assays. Peripheral blood was extracted for DNA using a Qiagen QIAamp DNA Blood Maxi kit, according to manufacturer’s instructions. The whole blood DNA samples were frozen at -80°C prior to extraction.

### Methylation data preprocessing

Genome-wide DNA methylation was measured using the Illumina EPIC array. In general, we preprocessed methylation data using an R package meffil^20^ based preprocessing pipeline (https://github.com/perishky/meffil/wiki).

We read and preprocessed blood DNA methylation data using the function meffil.qc, and removed ethnicity outliers, hip samples, samples with > 10% undetected (detection pvalue > 0.01) methylation values, sex outliers (>5 * sd), methylated/unmethylated signal outliers (> 3 * sd) and control probe signal outlier (>5 * sd). We then applied the same procedure (same R functions and thresholds) on DNA methylation samples from fat pad samples. Finally, we normalised methylation samples of all tissues together by applying meffil function meffil.normalize.quantiles (using 16 principal components) and meffil.normalize.samples.

We removed methylation sites with more than 10% of samples low bead number (< 3) or undetected methylation values (detection p < 0.01), non-autosomal methylation sites, methylation sites of cross-reactive probes and in close proximity (within 10 base pairs) to common SNPs (MAF > 0.05) in European population ^21–23^.

We converted initially generated beta values to Mvalues (beta2m function of R package lumi) ^24^ which we used for downstream analyses^25^. Per tissue, we further replaced strong outliers (>10 * sd from mean) with the methylation site-specific mean value. Based on a principal component analysis, we removed two outlier samples. The resulting fat pad methylation data comprised 780,177 methylation sites for 70 patients (46 women, 24 men, age 48-93 years, mean 71 years). For 58 of 70 patients, also methylation blood samples were available.

We used publicly available annotations (https://zwdzwd.github.io/InfiniumAnnotation/EPIC.hg38.manifest.tsv.gz and EPIC.hg38.manifest.gencode.v36.tsv.gz) to map probe identifier to the genomic location (hg38) and genes.

### Whole-genome sequencing data generation and preprocessing

Whole genome-sequencing (WGS) samples from the whole patient collection (n = 346 patients) were sequenced in two batches. In the first sequencing batch, genomic DNA was extracted from whole blood samples from 270 individuals (including 60 of 70 patients with QCed fat pad methylation data). In the second sequencing batch, genomic DNA from 73 samples from low-grade cartilage (including 8 of 70 patients with QCed fat pad methylation data) and three samples from synovium tissue was extracted. In both batches, DNA samples were subjected to standard Illumina paired-end DNA library construction, amplified, and subjected to DNA sequencing using the NovaSeq platform.

Generated CRAM files were input into samtools (samtools conda version 1.14) to create bam files. Subsequently, “bedtools bamtofastq” (bedtools conda version 2.30.0) was applied to obtain data in the fastq format. Per sequencing batch, variant calling was performed using the publicly available pipeline Sarek from nf-core (version 2.7.1, https://nf-co.re/sarek/2.7.1) with the additional options “-- tools HaplotypeCaller --generate_gvcf”. This uses the GATK Haplotypecaller (GATK v4.1.7.0) and generates g.vcf files. For the genome “GRCh38” was used. For the joint variant calling we adapted a publicly available pipeline (https://github.com/IARCbioinfo/gatk4-GenotypeGVCFs-nf) and used it with GATK (docker container broadinstitute/gatk:4.2.5.0). Reference files for GRCh38 were used from GATK.

For QC on the variant level, we applied Variant Quality Score Recalibration tool using a tranche threshold of 99.5% for SNPs and the recommended 99% for INDELs. For SNPs, this produces an expected false positive rate of 2.5% and an expected sensitivity of 97%.

For QC on the sample-level, we removed strong outlier het rate (two samples), and non-reference allele concordance when compared to directly typed genotype data using variants MAF > 0.01 (one sample), and one sample being a moderate outlier in sequencing depth as well as het rate. No additional sample was excluded solely based on Ti/Tv or singletons.

Furthermore, we removed one sex mismatch and two samples to avoid the inclusion of any sample pair with a relatedness > 0.2. We further excluded two ethnicity outliers identified in an ethnicity check-up using Ancestry and kinship toolkit (based on 1000G data from phase three; https://github.com/Illumina/akt/tree/master) ^26^. In total, we removed nine samples.

We excluded variants with MAF < 0.01, Hardy-Weinberg equilibrium p < 10^-5 and call rate <= 0.99. We then selected samples of individuals with matching fat pad methylation data (n = 68) and kept bi-allelic variants with MAF > 0.05. The resulting WGS data set used for the fat pad mQTL analysis comprised 68 samples and 6,395,994 variants.

### Comparing DNA methylation of blood and fat pad tissue

We integrated 70 fat pad and 58 blood samples in a principal component analysis (PCA) to investigate global epigenetic differences between these tissue types. We regressed out known technical batches (slide, row, clinical cohort) by applying Combat from the R package sva^27^ and performed PCA using prcomp function.

To compare methylation profiles on methylation site level, we performed differential methylation analysis between fat pad and blood samples paired from the same patients (n = 58). We performed linear modelling using the functions lmFit and eBayes from limma ^28^. We added the patient ID to ensure paired analysis design and included 19 surrogate variables (SVs) to account for technical variants. These SVs were estimated using the num.sv function from the sva package (‘be’-method) by protecting the tissue information. We highlighted methylation sites that exceed genome-wide significance threshold (Bonferroni correction with p < 6.41×10^-08 which corresponds to 0.05 / 780,177 methylation sites) with strong effect size (beta > 2).

### Methylation quantitative trait locus analysis

For the methylation quantitative trait locus (mQTL) analysis, we included whole-genome-sequencing data and fat pad methylation data matching from the same patients (n = 68). We included 6,395,994 bi-allelic genetic variants with a MAF > 0.05 among these 68 patients and set the cis-distance to 1Mb. We further normalised methylation levels using inverse-normal transformation per methylation site and estimated PEER factors^29^ (R package peer, default parameter setting) to infer hidden factors which we included to correct for technical variation. We performed cis-methylation QTL analysis using FastQTL (https://github.com/francois-a/fastqtl/)^30^. We first estimated nominal p values for every tested methylation site-variant pair using linear regression with the following model:

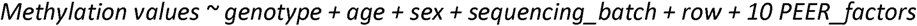

Here, *row* refers to the sample location on the Illumina EPIC array chip. The variable *sequencing_batch* accounts for WGS sequencing batches. Of 68 WGS samples, 60 and 8 were extracted in the first and second WGS sequencing batch, respectively (methods section “Whole-genome sequencing data generation and preprocessing”). To optimise the number of included PEER factors, we performed mQTL analysis with five, ten and 15 peer factors and chose the number that maximises detected mQTL targeted methylation site (5 Peer factors: 34,956 mQTL targeted methylation sites, 10 Peer factors: 35,948, 15 PEER factors: 35,808). Secondly, we applied an adaptive permutation scheme (implemented in FastQTL, parameter –permute 1000 10000) to estimate a q value and nominal p-value threshold per methylation site. Methylation sites with a q value < 5% Storey–Tibshirani FDR are regarded as mQTL targeted. For each mQTL-targeted methylation site, significant QTL were variants with a nominal p value below the nominal p value threshold for that methylation site.

### Colocalisation

We colocalised^31^ fat pad methylation QTL with GWAS signals for knee osteoarthritis and total knee replacement^2^. For this analysis, we applied the coloc.fast function (https://github.com/tobyjohnson/gtx/blob/526120435bb3e29c39fc71604eee03a371ec3753/R/coloc.R) using default settings. We performed colocalisation for mQTL-targeted methylation sites located within 100kb to an osteoarthritis GWAS index variant. For the colocalisation analysis, we included all variants that were included in the cis mQTL analysis for the tested methylation sites. We considered posterior probabilities (“PP4”) > 80% as indicator for colocalisation.

### Mendelian randomization

To estimate putative causal effects of QTL-targeted methylation sites in fat pad on osteoarthritis traits, we integrated the fat pad mQTL map with GWAS results for knee osteoarthritis and total knee replacement^2^. We applied two sample Mendelian randomization (MR) using the pipeline of the R package TwoSampleMR^32^. We considered methylation sites targeted by at least one mQTL. Per methylation site, we performed clumping (function clump_data, using the European reference panel and setting the R2 threshold to 0.01) to identify independent genetic variants which we included as instruments in the MR models. For methylation sites with one independent instrument, we applied the Wald-ratio, otherwise the inverse variance weighted method.

In total, we applied 64,898 MR models (32,448 and 32,450 for knee osteoarthritis and total knee replacement, respectively) to estimate the putative causal effect of 32,456 methylation sites. We applied the Bonferroni method to correct for multiple testing (p < 7.70×10^-07).

## Results

### Distinct epigenetic profiles in blood and fat pad adipocytes

We investigated global differences in the epigenetic profile between fat pad and peripheral blood samples for the first time. We performed PCA integrating infrapatellar fat pad samples from knee osteoarthritis patients (n = 70) and matched blood samples from a subset of these individuals (n = 58). We identified a separation of fat pad and blood samples along the first principal component, which was associated with tissue type (logistic regression p value: 2.7×10^-7, beta: -0.013, SE: 0.0026). This underlines the tissue-specificity of the epigenetic profile on a global level (Figure 1 A).

**Figure 1:**
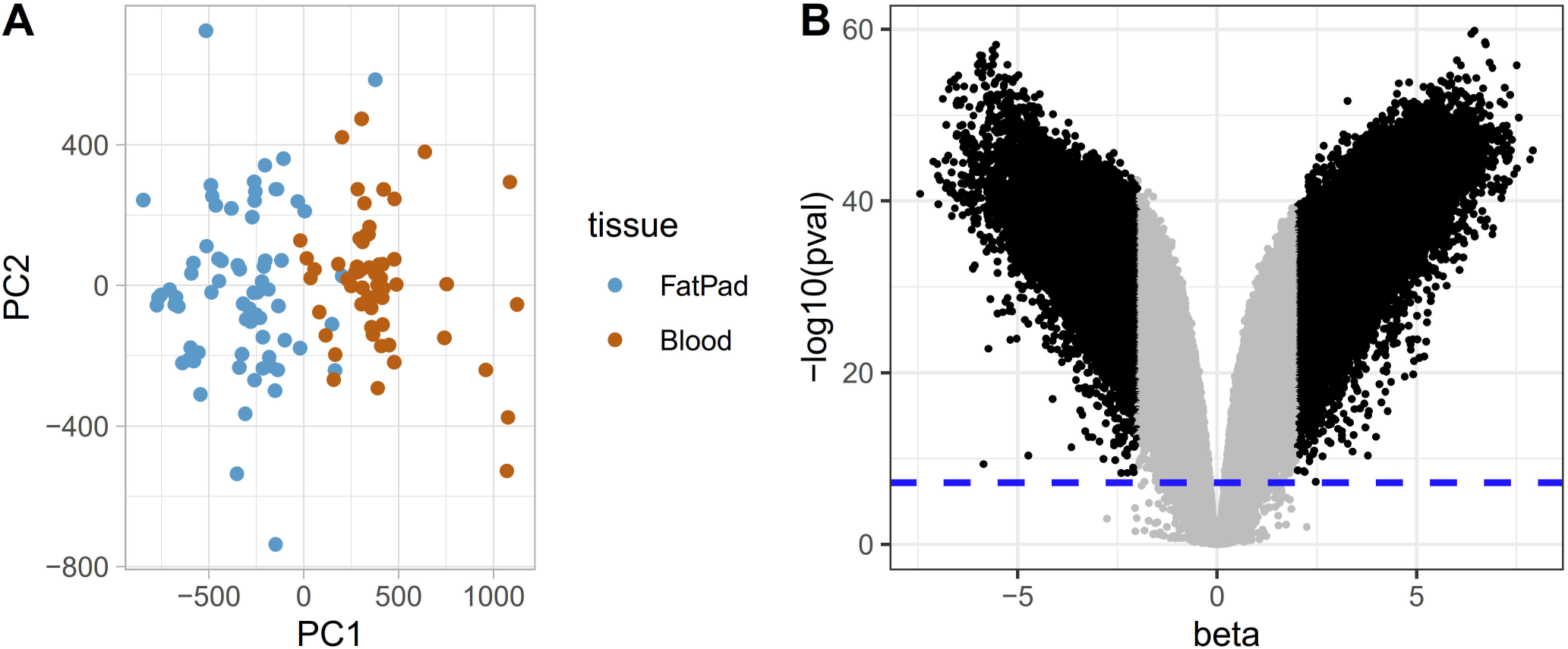
Distinct methylation profiles in blood and fat pad adipocytes. We investigated differences in the methylation profile between fat pad and blood. (A) On a global level, principal component analysis separates fat pad and blood samples along the first principal component. (B) On the methylation site level, a volcano plot demonstrates the multitude of differentially methylated sites. Sites with strong, differential methylation levels (beta > 2) exceeding the Bonferroni significance threshold (p < 6.41×10^-08, blue dashed line) are shown in black, otherwise in grey.

To characterise tissue-specificity on the methylation site level, we performed an epigenome-wide association study (EWAS) of matched fat pad and blood samples from the same patient (n = 58) and identified 84,973 (of 780,177 tested sites, 10.89 %) strongly differentially methylated sites (DMS) between fat pad and whole blood samples (p < 6.4×10^-08, beta > 2, Table S1). Of these, 33,391 and 51,582 showed hyper- and hypomethylation in fat pad tissue, respectively (Figure 1 B). Together, these results highlight extensive differences in the epigenetic profile of fat pad and peripheral blood.

### Genome-wide mQTL map in fat pad adipocytes

We performed cis-mQTL analysis to estimate genetic variants that are associated with methylation levels of nearby methylation sites (<= 1 Mb). We identified 35,948 mQTL-targeted methylation sites (Figure 2, Methods). Together, this constitutes the first genome-wide mQTL map of infrapatellar fat pad adipocytes in knee osteoarthritis. The full summary statistics are publicly available in the Musculoskeletal Knowledge Portal (http://mskkp.org).

**Figure 2:**
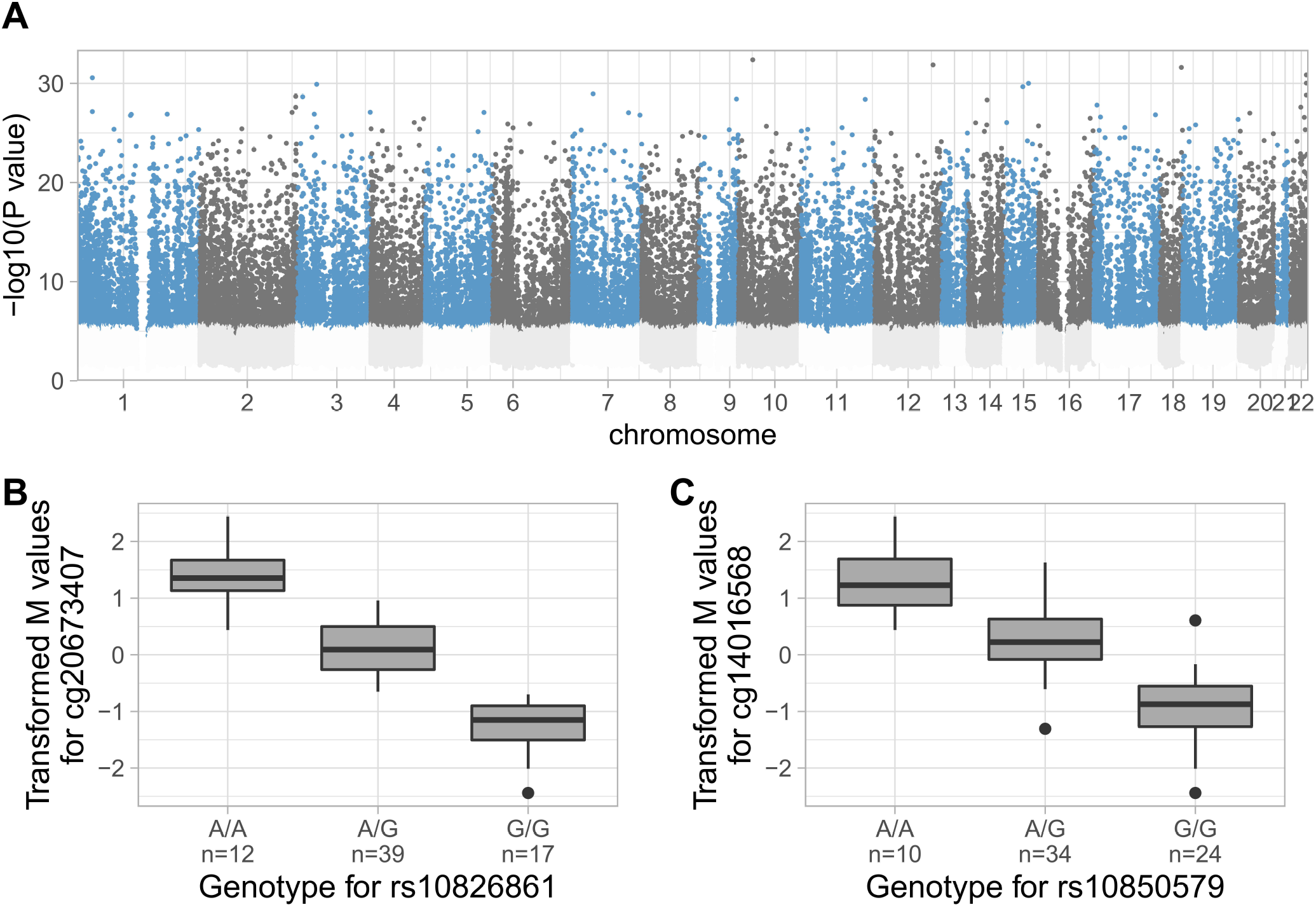
The mQTL map in fat pad adipocytes. (A) Manhattan plots depicting the negative log of the p value of the most significant association per methylation site across all variants. QTL targeted methylation sites are shown in blue or dark grey, otherwise in light grey. As examples, the boxplots illustrate the effect (B) of rs10826861 on cg20673407 (beta = -1.40, SE = 0.05, p = 4.15×10^-33) and (C) of rs10850579 on cg14016568 (beta = - 1.20, SE = 0.05, p = 1.10×10^-28). The boxplots represent 25th, 50th, and 75th percentiles, and whiskers extend to 1.5 times the interquartile range.

### Osteoarthritis GWAS signal resolution

Next, we integrated the newly-generated fat pad mQTL map with GWAS results of two osteoarthritis traits, namely knee osteoarthritis and total knee replacement^2^, to determine methylation sites with a putative causal role in osteoarthritis.

We applied colocalisation to estimate a probability for methylation mediating the osteoarthritis-promoting effect of risk variants. In total, we identified 16 methylation sites for which mQTL signals colocalised with 11 (of 25 tested, 44 %) GWAS signals (Posterior probability for colocalisation > 80%) (Table S2). For knee osteoarthritis, we resolved 9 (of 24 tested, 37.5 %) GWAS signals that colocalised with mQTL of 13 methylation sites (Figure 3A). Analogously, colocalising mQTL with GWAS results for total knee replacement resolved 5 (of 10 tested, 50%) GWAS signals and revealed 7 methylation sites with a potential causal role in osteoarthritis (Figure 3B).

**Figure 3:**
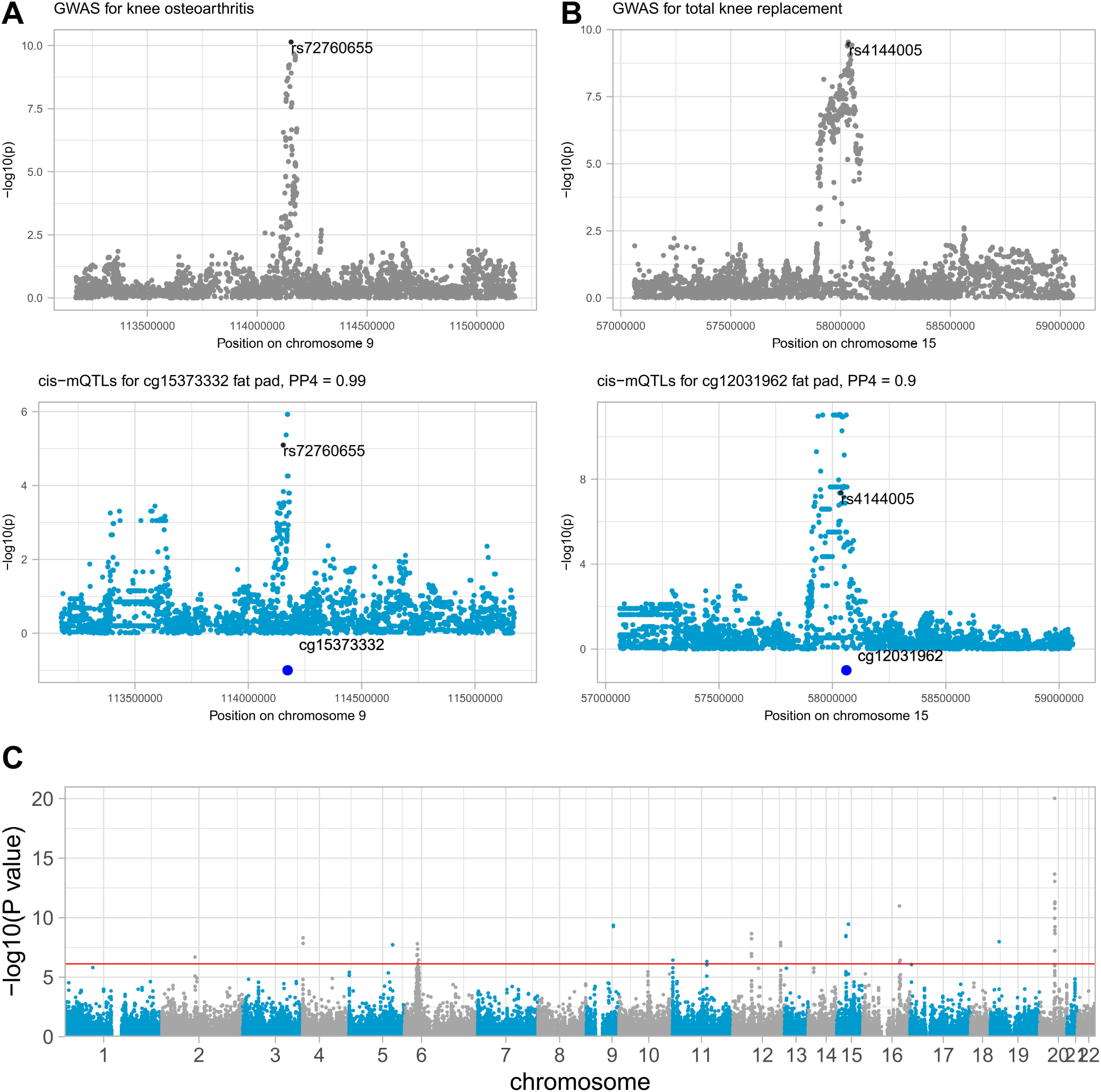
Osteoarthritis GWAS risk signals colocalise with mQTL. Two colocalisation events are exemplified in (A) and (B). In (A), we colocalised cis-mQTL for cg15373332 with a knee osteoarthritis GWAS signal (Posterior probability = 98.8 %). Similarly, (B) shows cis-mQTL for cg12031962 colocalising with a total knee replacement GWAS signal (Posterior probability = 89.8 %). (C) Manhattan plot depicting the Mendelian randomization p values to estimate the putative causal effects of methylation sites in fat pad on knee osteoarthritis or total knee replacement. The red line indicates genome-wide significance applying the Bonferroni correction (p < 8.31×10^-07).

Next, we performed causal inference analysis by applying two-sample Mendelian randomization (MR) to estimate the putative causal effect of methylation on osteoarthritis. In these MR models, we used mQTL as instruments as well as mQTL-targeted methylation sites and osteoarthritis as exposure and outcome, respectively (Method). Here, we detected 36 methylation sites with a putative role (p < 7.70×10^-07) in osteoarthritis (Figure 3C), in total (Table S3). For knee osteoarthritis, we identified 32 methylation sites, of which 15 and 17 revealed a link of hyper- and hypomethylation with osteoarthritis, respectively. For total knee replacement, we identified 15 methylation sites with a putative causal role (9 and 6 showing hyper- and hypomethylation in osteoarthritis, respectively). Eleven methylation sites were identified in both osteoarthritis-relevant traits, for which the direction of effect was concordant.

MR and colocalisation identified 37 putative causal methylation sites, in total. Of these, 15 were identified in both approaches, thus providing two lines of evidence for their respective causal involvement (Table S4). Together, colocalisation and MR results suggest that these methylation marks mediate the regulation of genetic risk variants on effector genes in fat pad.

Annotated genes of the identified 37 methylation sites have been previously linked to osteoarthritis using causal approaches on genome-wide mQTL maps of cartilage or synovium. This includes *WWP2* (annotated to fat pad relevant methylation site cg04703221), a chondrocyte regulator^33^ for which methylation has been causally linked to osteoarthritis in low disease-grade cartilage and synovium^7^. *ALDH1A2* (cg12031962, cg12031962 and cg08668585) has also previously been linked to osteoarthritis at the methylation^7^ (in low-and high-grade osteoarthritis cartilage as well as synovium) as well as expression^6^ (low-grade osteoarthritis cartilage) levels. Furthermore, we identified osteoarthritis-linked methylation in the collagen type *COL27A1*(cg21771125).

We also identified likely effector genes that were not previously resolved in molecular QTL maps of primary osteoarthritis cartilage and synovium^6,7^ including *USP8* (cg01701297 and cg05456662; involved in cell proliferation), *TSKU (*cg17107561; encodes development-linked extracellular matrix protein) and *FER1L4* (cg14387502 cg05220160; involved in plasma membrane organization) which can be linked to osteoarthritis-relevant mechanisms (Discussion). Together, integrating the fat pad mQTL profile with osteoarthritis GWAS results using colocalisation and MR identified 37 methylation sites with a potential causal involvement in osteoarthritis in fat pad tissue.

## Discussion

Osteoarthritis is a common joint disorder with a polygenic architecture. Genome-wide molecular profiles of affected primary tissues remain understudied and excluded from large molecular data resources, such as GTEx^34^, ENCODE^35^ and RoadMap^35^. In this study, we characterised the first epigenome-wide profile of osteoarthritis-affected infrapatellar fat pad. We identify extensive differences from the epigenetic profile of peripheral blood, generate the first genome-wide mQTL map in fat pad, and identify methylation sites with a likely causal role in osteoarthritis development and progression.

Comparing fat pad and blood methylation profiles reveals abundant epigenetic differences underlining the epigenetic tissue-specificity of blood and joint tissues, thus highlighting the necessity to investigate disease-affected tissues.

We present the first genome-wide mQTL map for osteoarthritis-affected infrapatellar fat pad. Colocalising this mQTL map with osteoarthritis GWAS results resolved eleven genetic osteoarthritis risk signals, thus providing evidence for methylation mediating the genetic effect of these GWAS signals on osteoarthritis in fat pad.

We supplemented these causal insights using MR and, together with colocalisation, identified 37 methylation sites with a putative causal role in osteoarthritis in fat pad. Some methylation sites were close to genes (such as *WWP2, ALDH1A2*, and *COL27A1*) that have been previously causally linked to osteoarthritis using genome-wide molecular QTL maps of other primary joint tissues^6,7^, suggesting a disease-relevant role across joint tissues.

We also identify genes that have not been previously resolved in molecular QTL maps of primary osteoarthritis tissues^6,7^ such as *USP8, TSKU* and *FER1L4*. USP8 is involved in epidermal growth factor receptor regulation^36^, a receptor linked to angiogenic and inflammatory mechanisms. *TSKU* is a regulator of Wnt signaling, a pathway which has been consistently linked to osteoarthritis across tissues, e.g. in cartilage, synovium and subchondral bone^37^. *FER1L4* regulates inflammatory factor IL-6 in osteoarthritis-affected cartilage^38^ and is linked to VEGF, an osteoarthritis-linked angiogenic factor^39^. Together, we have identified genes linked to processes that are observed in osteoarthritis-affected fat pad, such as inflammation or vascularization^40^, and suggest an involvement of the detected methylation sites in disease-related alterations.

Our findings highlight differences in the epigenetic profile of fat pad tissue and blood and identify methylation sites that likely exert the effect of GWAS risk signals in fat pad, shedding light on the mechanistically relevant role of fat pad methylation in osteoarthritis.

## Data Availability

Full summary statistics will be made openly available through the MSK portal (http://mskkp.org) upon manuscript acceptance.

## Acknowledgements

We acknowledge the technical support of Core Facility Genomics at Helmholtz Munich. We thank Dr. Inti Alberto de la Rosa Velasquez, Dr. Peter Lichtner, Susanne Wittmann and Dr. Thomas Walzthöni for help with DNA methylation and whole-genome sequencing data generation as well as whole-genome sequencing data preprocessing.

## Author contributions

Study design: E.Z., J.M.W.; Clinical collection: J.M.W., D.S.; WGS data preprocessing: A.G., Y.C.P.; Data analysis: P.K.; Interpretation of results: P.K., E.Z.; Manuscript drafting: P.K., E.Z.; Manuscript reviewing and editing: P.K., E.Z., J.M.W., D.S.

## Data and code availability

Full summary statistics will be made openly available through the MSK portal (http://mskkp.org) upon manuscript acceptance. All software used in this study is available from free repositories or manufacturers as referenced in the Subjects and methods section.

